# Effect of common maintenance drugs on the risk and severity of COVID-19 in elderly patients

**DOI:** 10.1101/2021.09.28.21264186

**Authors:** Kin Wah Fung, Seo H. Baik, Fitsum Baye, Zhaonian Zheng, Vojtech Huser, Clement J. McDonald

## Abstract

**Background:** Maintenance drugs are used to treat chronic conditions. Several classes of maintenance drugs have attracted attention because of their potential to affect susceptibility to and severity of COVID-19.

**Methods:** Using claims data on 20% random sample of Part D Medicare enrollees from April to December 2020, we identified patients diagnosed with COVID-19. Using a nested case-control design, non-COVID-19 controls were identified by 1:5 matching on age, race, sex, dual-eligibility status and geographical region. We identified usage of angiotensin-converting enzyme inhibitors (ACEI), angiotensin-receptor blockers (ARB), warfarin, direct factor Xa inhibitors, clopidogrel, famotidine and hydroxychloroquine based on Medicare prescription claims data. Using extended Cox regression models with time-varying propensity score adjustment we examined the independent effect of each study drug on contracting COVID-19. For severity of COVID-19, we performed extended Cox regressions on all COVID-19 patients, using COVID-19-related hospitalization and all-cause mortality as outcomes. Covariates included gender, age, race, geographic region, low-income indicator and co-morbidities. To compensate for indication bias related to the use of hydroxychloroquine for the prophylaxis or treatment of COVID-19, we censored patients who only started on hydroxychloroquine in 2020.

**Results:** Up to December 2020, our sample contained 374,229 Medicare patients over 65 who were diagnosed with COVID-19. Among the COVID-19 patients, 209,208 (55.9%) were on at least one study drug. The three most common study drugs were ACEI 97,872 (26.1%), ARB 83,329 (22.3%) and clopidogrel 38,203 (10.2%). Current users of ACEI, ARB, warfarin, direct factor Xa inhibitor and clopidogrel were associated with reduced risk of getting COVID-19 (3-13%), and reduced risk of dying after a COVID-19 diagnosis (8-19%). Famotidine did not show consistent significant effects. Hydroxychloroquine did not show significant effects after censoring of recent starters.

**Conclusions:** Maintenance use of ACEI, ARB, warfarin, direct factor Xa inhibitor and clopidogrel was associated with reduction in risk of acquiring COVID-19 and dying from it.

## 1. Background and significance

Maintenance drugs are indicated for chronic conditions, taken indefinitely on regular, usually daily basis. Reports have attributed protective or aggravating effects of some maintenance drugs on the likelihood and/or severity of SARS-CoV-2 infection. These include angiotensin-converting enzyme inhibitors (ACEIs), angiotensin-receptor blockers (ARBs), antithrombotic agents, hydroxychloroquine and famotidine.

ACEI and ARB attracted early attention because they increase the expression of angiotensin-converting enzyme 2 (ACE2), [1, 2] the cellular doorway for SARS-CoV-2. [3, 4] There are concerns that their use could *increase* the likelihood, or the severity, of a COVID-19 infection. However, most studies published to-date show no such increases, [5-9] and some actually suggest that they *might decrease* COVID-19 severity. [10, 11] Due to the potential beneficial effect, several ongoing prospective trials are testing whether initiating ACEI or ARB treatment for patients after they are diagnosed with COVID-19 would confer any benefits. [12-14]

Given the thrombotic propensity of COVID-19, [15-19] some think that antithrombotics could reduce the severity of COVID-19, and accordingly, anticoagulants are now recommended for hospitalized and high risk patients, [20] though results of studies of maintenance antithrombotics have been mixed. [21-25] Famotidine has attracted attention because it has anti-retroviral activity and it improved COVID-19 outcomes in small studies. [26] [27, 28] Hydroxychloroquine was considered as treatment option for COVID-19 because of its immune modulation and anti-SARS-CoV-2 activity in vitro, [29-31] and reported benefit in one clinical study. [32]

Almost all of the primary studies on the effect of these maintenance drugs have been small (tens or hundreds of COVID-19 patients who were also on the drugs) and would mostly likely lack statistical power to see important associations.

In the U.S., the Virtual Research Data Center (VRDC) [33] of the Centers for Medicare and Medicaid Services (CMS) carries de-identified data which include complete drug prescription, diagnoses, encounters as well as geographic, socio-demographic and vital status information for most (>93%) of the 65 and older U.S. residents. Over 80% of COVID-19 deaths occur in this age group, according to the U.S. Centers for Disease Control and Prevention (CDC). Therefore, this would be a propitious population in which to explore the association of maintenance drugs with COVID-19 risks.

In this study, we used the VRDC data and extended Cox regression analysis to examine associations between the above seven drug types and the occurrence of three outcomes: 1) COVID-19 infection, 2) COVID-19 hospitalization, and 3) death after a COVID19 diagnosis.

## 2. Materials and methods

### 2.1 Study population and case definition

VRDC provided us with a 20% random sample of all Medicare Part D enrollees. This set included 374,299 patients 65 or above, who had at least one record of the COVID-19 specific ICD10-CM diagnosis code of U07.1 between April 1 and December 31, 2020. We used both inpatient and outpatient claims and Medicare’s vital status to identify COVID-19 cases and the outcome events. We only counted COVID-19 cases occurring on or after April 1, 2020, when the specific code for COVID-19 first became available. We stopped accruing cases after December 31, 2020 because of potential incompleteness of data due to the time lag (at least 3-4 months) between data capture and availability through VRDC. This study was declared not human subject research by the Office of Human Research Protection at the National Institutes of Health and by the CMS’s Privacy Board.

### 2.2 Drugs, exposure definition and comorbidities

We identified all study drug preparations that were available in the U.S. market. For drug classes we identified the class members through Anatomical Therapeutic Chemical (ATC) classification codes and used generic drug names to find them in the CMS data. Our seven study drugs/drug classes were ACEI, ARB, warfarin, direct factor Xa inhibitors, clopidogrel, famotidine and hydroxychloroquine (see Supplementary Table 1 for full list of drugs and prescription frequencies). Note that hydroxychloroquine was a class which also included two other aminoquinolines (chloroquine and primaquine), but hydroxychloroquine accounted for 99.7% of prescriptions.

We assumed that patients were on a given study drug during the window from the prescription dispensing date to 30 days after the end-of-supply day (we called this period the “current use period”). We added the 30-day buffer after the end-of-supply day because of the common behavior of drug stockpiling – patients maintaining a stock of drugs so that they will not run out immediately in case refill is interrupted. Drug use status was treated as a time-varying covariate in the Cox regression model. We defined a patient to be a current user if an outcome event fell within a current use period, former user if the event fell outside the current use period, and never user if they never had a prescription for that drug. Our primary analysis compared current users with never users as control. In a supplementary analysis, we used former users as control to see if that would give different results.

In the first few months of the pandemic, there were media reports of the potential beneficial or harmful effects of some of the study drugs on COVID-19. Publicity about the good effects of hydroxychloroquine was particularly rife, which could have led people to start taking it because of symptoms, or fear, of COVID-19. We looked at the trend of the usage of the study drugs, starting from January, 2019, to identify abnormal patterns that could be related to COVID-19. To study the potential effect of COVID-19 affecting the use of drugs (“reverse causality”), we did separate analyses with special treatment of patients who were only recently started on a study drug (see section 2.3 below).

Medicare specifies the onset of 67 chronic conditions by algorithm and we followed the algorithm to define the occurrence and onset date of each condition. [34] To adjust for illness burden in our analysis we included 57 Medicare chronic conditions with >1% prevalence in the Master Beneficiary Summary File.

### 2.3 Statistical analysis and covariates

We excluded patients who were not enrolled in Parts A (hospital) and B (medical) to ensure complete capture of hospitalization and encounter diagnoses. We considered the effect of the study drugs on three outcomes: 1) the risk of acquiring a COVID-19 diagnosis, 2) the risk of COVID-19 hospitalization, and 3) the risk of death after being diagnosed with COVID-19. For the first outcome (diagnosis of COVID-19), we used a nested case-control analysis,[35] where each index COVID-19 patient was matched to patients with no COVID-19 diagnosis up to the date COVID-19 was diagnosed in the index patient. Each index case was matched to five controls on age in years, race, sex, dual-eligibility status and five regions of residence (Northeast, Midwest, South, West and others) at the time of the diagnosis of COVID-19. All cases and control were followed from January 1, 2020 until COVID-19 diagnosis, death, disenrollment from Medicare Parts A/B/D or December 31, 2020, whichever came first.

For the analyses of the second and third outcomes (hospitalization and death), we included only patients diagnosed with COVID-19. For the hospitalization outcome, we followed all patients from COVID-19 diagnosis until COVID-19-related hospitalization, or the other censoring points as described for the first outcome. For the death outcome, we did the same, swapping hospitalization for death.

We approximated patient’s income level using the monthly indicators of dual-eligibility and low-income subsidy (LIS), which divided patients into three income groups: 1) dual-eligible: income ≤ 135% federal poverty line (FPL), 2) non-dual LIS: income > 135% and ≤ 150% FPL, and 3) non-dual no LIS: income > 150% FPL. To explore the effect of each study drug, we employed extended Cox regression analysis with days-on-study as time scale. We included age, gender, race, regions of residence, Medicare insurance type (Advantage or fee-for-service) and the degree of LIS as covariates. We also included binary flags for each of the 57 Medicare chronic conditions to control for the effect of co-morbidities. In order to protect against the immortal time bias and violation of proportional hazard assumption,[36] we treated the use of each of the study drugs and almost all covariates as time-varying covariates. Only age, sex, race and regions of residence were time-fixed, and their values were defined as of their values on January 1, 2020. Since the disease covariates were chronic diseases, we considered them as always present after their onset and always absent before then. The values of all time-varying covariates were reset at the time of each event in the Cox regression. We used Efron’s adjustment for tied events.

To mitigate the potential of COVID-19 influencing the use of drugs, we did a separate Cox regression in which patients who only started a study drug in 2020 (they had never been on the drug before January 1,2020) were censored when they first started the drug (“recent starter censoring”). We compared the results with and without such censoring.

In order to mitigate selection bias toward use of the study drugs, we developed time-varying propensity scores (PS) [37] separately for each study drug using logistic regressions. The PS was the likelihood of receiving a study drug, conditional on patient’s characteristics (demographics, socioeconomics, and presence of the chronic conditions). We iteratively estimated the PSs every month among the patients who remained in follow-up, considering all covariates that preceded the end of a given monthly cycle, [38] and ran all Cox regression analyses with time-varying PSs as additional adjustments.

## 3. Results

### 3.1 Study population

In our 20% random sample of Part D enrollees, 374,299 Medicare beneficiaries aged 65 and above were diagnosed with COVID-19 between April 1 and December 31, 2020, among which 65,108 (17.4%) died (Table 1 and Consort diagram in supplementary material). Over the same period, CDC registered a national total of 19,852,636 COVID-19 cases and 350,510 deaths.[39] In the CDC statistics, even though patients ≥65 accounted for only 13.9% of cases, they accounted for 80.5% of mortalities. Therefore, our study population represents patients most at risk for COVID-19 death. Projecting from the CDC statistics, a 20% random sample of patients 65 and above would have 551,903 COVID-19 cases and 56,432 mortalities (Table 1). Therefore, our study population captured 67.8% of COVID-19 cases nationally. Our mortality number is 15% higher than the projected national count. Our death numbers were higher because Medicare data did not distinguish between causes of death. It is possible that some mortalities were not related to COVID-19. However, most of the mortalities occurred within a short time after the COVID-19 diagnosis (median 14 days, inter-quartile range 6 – 36 days). Compared with national statistics, the Medicare population was over-represented in females, whites and under-represented in Hispanics. Geographically, there was over-representation of the North-East region.

**Table 1.** Study population compared to U.S. national statistics, April to December 2020 (SRS – simple random sample; CDC - Centers for Disease Control and Prevention)

|  | **Medicare 20% SRS Age 65+ population**<br>**(Apr-Dec 2020)** |  | **National statistics from CDC**<br>**(Apr-Dec 2020)** |  |
| --- | --- | --- | --- | --- |
|  | ***Covid-19 cases (%)*** | ***Mortality (%)*** | ***Covid-19 cases*** | ***Mortality*** |
| Total | 374,299(100) | 65,108(100) | 551,903* | 56,432* |
| Female | 225,585(60.3) | 36,093(55.0) | 52.2% | 45.7% |
| ***Race*** |  |  |  |  |
| White | 284,707(76.1) | 46,595(71.6) | 50.0% | 58.6% |
| Black | 38,419(10.3) | 8,702(13.4) | 11.0% | 13.6% |
| Hispanic | 31,981(8.5) | 6,401(9.8) | 29.2% | 18.9% |
| Asian | 9,961(2.7) | 2,032(3.1) | 3.3% | 4.0% |
| Other | 9,231(2.5) | 1,378(2.1) | 6.6% | 5.0% |
| ***Geographic region*** |  |  |  |  |
| North East | 82,281(22.0) | 16,292(25.0) | 17.15% | 23.45% |
| Midwest | 90,921(24.3) | 15,449(23.7) | 21.76% | 20.18% |
| South | 139,567(37.3) | 24,248(37.2) | 38.95% | 36.91% |
| West | 60,830(16.3) | 9,046(13.9) | 21.71% | 19.03% |

\*The national case and mortality counts are extrapolations from the CDC data. They only include patients aged 65+, and reduced to 20% to be comparable with our 20% random sample

### 3.2 Drug exposure and matching

Table 2 shows the breakdown of drug usage (all patients who had prescription for the drug during the follow up period, starting on January 1, 2020) among the COVID-19 patients and controls. Overall, 209,208 (55.9%) cases and 906,326 (48.6%) controls had prescriptions for at least one study drug. For all drugs, the usage was significantly higher among COVID-19 patients than controls. The unmatched rate (less than five controls found) was 0.4%.

**Table 2.** Characteristics of Covid-19 patients and matched controls

|  | **Covid-19 patients (%)** | **Control (%)** | **(Control -Covid) Difference (95%CI)** | **PVALUE** |
| --- | --- | --- | --- | --- |
| **Drug exposure** |  |  |  |  |
| ACE Inhibitor | 97,872(26.1) | 451,115(24.2) | -1.9(-2.1,-1.8) | 0.000 |
| ARB | 83,329(22.3) | 383,276(20.5) | -1.8(-1.9,-1.7) | 0.000 |
| Warfarin | 11,757(3.1) | 41,722(2.2) | -0.9(-1.0,-0.8) | 0.000 |
| Direct Factor Xa Inhibitor | 14,148(3.8) | 44,706(2.4) | -1.4(-1.4,-1.3) | 0.000 |
| Clopidogrel | 38,203(10.2) | 120,957(6.5) | -3.7(-3.8,-3.6) | 0.000 |
| Hydroxychloroquine | 2,879(0.8) | 10,910(0.6) | -0.2(-0.2,-0.1) | 0.000 |
| Famotidine | 13,147(3.5) | 30,539(1.6) | -1.9(-2.0,-1.8) | 0.000 |
| Any drug | 209,208(55.9) | 906,326(48.6) | -7.3(-7.5,-7.2) | 0.000 |
| No Rx | 165,091(44.1) | 960,250(51.4) | 7.3(7.2,7.5) | 0.000 |
| **Age** |  |  |  |  |
| 65 - 69 | 37,859(10.1) | 189,150(10.1) | 0.0(-0.1,0.1) | 0.839 |
| 70 - 74 | 89,538(23.9) | 447,650(24.0) | 0.0(-0.1,0.2) | 0.582 |
| 75 - 79 | 77,520(20.7) | 387,477(20.8) | 0.0(-0.1,0.2) | 0.662 |
| 80 - 84 | 64,951(17.4) | 324,637(17.4) | 0.0(-0.1,0.2) | 0.704 |
| 85 + | 104,138(27.8) | 517,662(27.7) | -0.1(-0.3,0.0) | 0.168 |
| **Region** |  |  |  |  |
| MIDWEST | 90,840(24.3) | 453,053(24.3) | -0.0(-0.2,0.1) | 0.830 |
| NORTHEAST | 82,225(22.0) | 410,408(22.0) | 0.0(-0.1,0.1) | 0.976 |
| SOUTH | 139,505(37.3) | 696,758(37.3) | 0.0(-0.1,0.2) | 0.747 |
| WEST | 60,761(16.2) | 303,161(16.2) | -0.0(-0.1,0.1) | 0.946 |
| OTHER | 675(0.2) | 3,196(0.2) | -0.0(-0.0,0.0) | 0.213 |
| Unmatched | 293 (< 0.1) | - |  |  |
| **Total** | **374,299 (100)** | **1,866,576 (100)** |  |  |
| Number of patients with < 5 matches | 1446 (0.4) |  |  |  |

As for abnormal patterns of drug usage related to COVID-19, we detected a sharp rise in the use of hydroxychloroquine in March and April 2020 (Supplementary Figure 2), which was not seen in other study drugs. FDA granted emergency use authorization for hydroxychloroquine for COVID-19 treatment on March 28, and the percentage of COVID-19 patients on hydroxychloroquine surged to over 0.8% compared to the baseline of 0.3% before the pandemic. There was a similar but smaller rise in non-COVID-19 patients, showing that some patients might be taking hydroxychloroquine for prophylaxis or suspicion of COVID-19. After FDA revoked the emergency use authorization on June 15, the usage of hydroxychloroquine began to drop, but remained slightly above the baseline before the pandemic.

### 3.3 R*isk of being diagnosed with COVID-19*

The total number of patients followed (cases and controls) was about 2.2 million, and around 350,000 ended up with the diagnosis of COVID-19 (Table 3). We did separate analyses with and without recent starter censoring for all study drugs. Only hydroxychloroquine exhibited significantly different results with censoring. We report the results with recent starter censoring for hydroxychloroquine as the main results in table 3. The results for hydroxychloroquine without such censoring are shown for comparison.

**Table 3.**
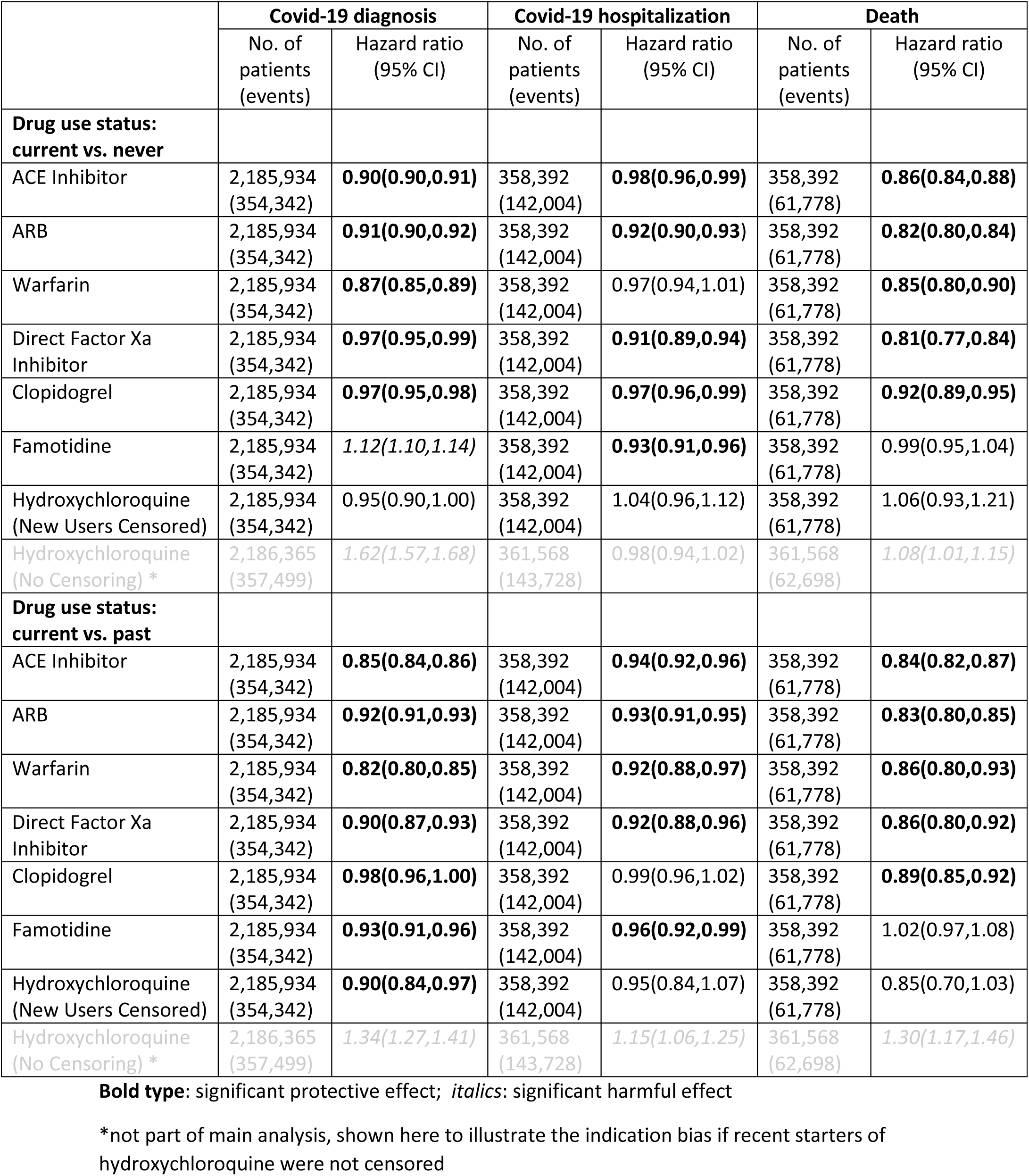
Effect of drug use on Covid-19 diagnosis, Covid-19 hospitalization and death

|  | Covid-19 diagnosis |  | Covid-19 hospitalization |  | Death |  |
| --- | --- | --- | --- | --- | --- | --- |
|  | No. of patients (events) | Hazard ratio (95% CI) | No. of patients (events) | Hazard ratio (95% CI) | No. of patients (events) | Hazard ratio (95% CI) |
| **Drug use status: current vs. never** |  |  |  |  |  |  |
| ACE Inhibitor | 2,185,934 (354,342) | **0.90(0.90,0.91)** | 358,392 (142,004) | **0.98(0.96,0.99)** | 358,392 (61,778) | **0.86(0.84,0.88)** |
| ARB | 2,185,934 (354,342) | **0.91(0.90,0.92)** | 358,392 (142,004) | **0.92(0.90,0.93)** | 358,392 (61,778) | **0.82(0.80,0.84)** |
| Warfarin | 2,185,934 (354,342) | **0.87(0.85,0.89)** | 358,392 (142,004) | 0.97(0.94,1.01) | 358,392 (61,778) | **0.85(0.80,0.90)** |
| Direct Factor Xa Inhibitor | 2,185,934 (354,342) | **0.97(0.95,0.99)** | 358,392 (142,004) | **0.91(0.89,0.94)** | 358,392 (61,778) | **0.81(0.77,0.84)** |
| Clopidogrel | 2,185,934 (354,342) | **0.97(0.95,0.98)** | 358,392 (142,004) | **0.97(0.96,0.99)** | 358,392 (61,778) | **0.92(0.89,0.95)** |
| Famotidine | 2,185,934 (354,342) | *1.12(1.10,1.14)* | 358,392 (142,004) | **0.93(0.91,0.96)** | 358,392 (61,778) | 0.99(0.95,1.04) |
| Hydroxychloroquine (New Users Censored) | 2,185,934 (354,342) | 0.95(0.90,1.00) | 358,392 (142,004) | 1.04(0.96,1.12) | 358,392 (61,778) | 1.06(0.93,1.21) |
| Hydroxychloroquine (No Censoring) * | 2,186,365 (357,499) | *1.62(1.57,1.68)* | 361,568 (143,728) | 0.98(0.94,1.02) | 361,568 (62,698) | *1.08(1.01,1.15)* |
| **Drug use status: current vs. past** |  |  |  |  |  |  |
| ACE Inhibitor | 2,185,934 (354,342) | **0.85(0.84,0.86)** | 358,392 (142,004) | **0.94(0.92,0.96)** | 358,392 (61,778) | **0.84(0.82,0.87)** |
| ARB | 2,185,934 (354,342) | **0.92(0.91,0.93)** | 358,392 (142,004) | **0.93(0.91,0.95)** | 358,392 (61,778) | **0.83(0.80,0.85)** |
| Warfarin | 2,185,934 (354,342) | **0.82(0.80,0.85)** | 358,392 (142,004) | **0.92(0.88,0.97)** | 358,392 (61,778) | **0.86(0.80,0.93)** |
| Direct Factor Xa Inhibitor | 2,185,934 (354,342) | **0.90(0.87,0.93)** | 358,392 (142,004) | **0.92(0.88,0.96)** | 358,392 (61,778) | **0.86(0.80,0.92)** |
| Clopidogrel | 2,185,934 (354,342) | **0.98(0.96,1.00)** | 358,392 (142,004) | 0.99(0.96,1.02) | 358,392 (61,778) | **0.89(0.85,0.92)** |
| Famotidine | 2,185,934 (354,342) | **0.93(0.91,0.96)** | 358,392 (142,004) | **0.96(0.92,0.99)** | 358,392 (61,778) | 1.02(0.97,1.08) |
| Hydroxychloroquine (New Users Censored) | 2,185,934 (354,342) | **0.90(0.84,0.97)** | 358,392 (142,004) | 0.95(0.84,1.07) | 358,392 (61,778) | 0.85(0.70,1.03) |
| Hydroxychloroquine (No Censoring) * | 2,186,365 (357,499) | *1.34(1.27,1.41)* | 361,568 (143,728) | *1.15(1.06,1.25)* | 361,568 (62,698) | *1.30(1.17,1.46)* |

**Bold type:** significant protective effect; *italics:* significant harmful effect
\*not part of main analysis, shown here to illustrate the indication bias if recent starters of hydroxychloroquine were not censored

Compared to patients who never used the drug, current user of ACEI, ARB, warfarin, direct factor Xa inhibitor and clopidogrel was associated with decreased risk of contracting COVID-19, ranging from 3% reduction (direct factor Xa inhibitor and clopidogrel) to 13% reduction (warfarin). For famotidine, there was a 12% increase in risk. For hydroxychloroquine, with the proper censoring of recent starters, there was no effect on the risk of being diagnosed with COVID-19. However, without censoring, current users would appear to be associated with an increased risk of getting COVID-19 compared to never users (62% increase).

The results for the non-drug covariates are shown in Table 4. Compared with the youngest group of patients in our population (65-69), 70-74 year-olds were associated with a greater risk (5% increase) of a COVID-19 diagnosis. However, the risk of COVID-19 diagnosis dropped monotonically with age above 75, with 22% reduced risk for patients over 85. Female experienced a 17% risk reduction compared to males. The risk of COVID-19 diagnosis was higher in all race groups compared to whites, 28% higher for Asians, 39% for Blacks and 70% for Hispanics. The comorbidities associated with the greatest increase in risk were dementia (104%), hypertension (78%) and schizophrenia (51%).

**Table 4.** Effect of non-drug covariates on Covid-19 diagnosis, Covid-19 hospitalization and death

| Co-variate | Reference | COVID-19 diagnosis | COVID hospitalization | Death |
| --- | --- | --- | --- | --- |
|  |  | Hazard ratio (95% CI) |  |  |
| **Demographics** |  |  |  |  |
| 70-74 | 65-69 | 1.05(1.03,1.06) | 1.11(1.09,1.13) | 1.12(1.08,1.16) |
| 75-79 | 65-69 | **0.92(0.90,0.93)** | 1.43(1.40,1.46) | 1.55(1.49,1.61) |
| 80-84 | 65-69 | **0.81(0.80,0.82)** | 1.66(1.62,1.70) | 1.98(1.90,2.06) |
| >85 | 65-69 | **0.78(0.77,0.79)** | 1.97(1.92,2.02) | 3.12(2.99,3.25) |
| Female | Male | **0.83(0.82,0.85)** | **0.79(0.78,0.81)** | **0.66(0.64,0.68)** |
| Black | white | 1.39(1.37,1.41) | 0.99(0.97,1.01) | 1.04(1.01,1.08) |
| Hispanic | white | 1.70(1.68,1.73) | 1.15(1.13,1.18) | 1.23(1.20,1.27) |
| Asian | white | 1.28(1.23,1.33) | 1.05(1.00,1.10) | 1.36(1.28,1.46) |
| Other | white | 1.14(1.11,1.16) | 1.07(1.03,1.11) | 1.29(1.22,1.36) |
| Dual | Non-dual<br>No LIS | **0.45(0.45,0.46)** | 1.17(1.15,1.20) | **0.72(0.70,0.75)** |
| Non-dual LIS | Non-dual<br>No LIS | **0.78(0.76,0.79)** | 1.47(1.42,1.52) | 3.15(3.04,3.27) |
| Midwest | Northeast | 1.04(1.03,1.05) | 1.38(1.36,1.41) | 1.05(1.02,1.08) |
| South | Northeast | 1.41(1.39,1.43) | 1.20(1.18,1.22) | **0.96(0.94,0.98)** |
| West | Northeast | **0.69(0.63,0.76)** | 1.16(1.13,1.18) | **0.89(0.87,0.92)** |
| Other | Northeast | **0.94(0.93,0.95)** | **0.41(0.33,0.51)** | **0.58(0.45,0.76)** |
| **Comorbidities** |  |  |  |  |
| ESRD | No | 1.35(1.30,1.41) | **0.86(0.82,0.89)** | 1.63(1.53,1.73) |
| AMI | No | **0.91(0.89,0.92)** | 1.18(1.16,1.21) | 1.40(1.36,1.44) |
| Atrial Fibrillation | No | **0.45(0.44,0.46)** | **0.95(0.92,0.97)** | 1.16(1.11,1.20) |
| Cataract | No | 1.01(1.00,1.02) | **0.84(0.82,0.85)** | **0.82(0.80,0.84)** |
| Chronic Kidney Disease | No | 1.07(1.06,1.08) | 1.33(1.32,1.35) | 1.58(1.55,1.62) |
| COPD | No | **0.93(0.92,0.94)** | **0.95(0.94,0.97)** | 1.07(1.05,1.09) |
| Heart Failure | No | **0.98(0.97,0.99)** | 1.02(1.01,1.03) | 1.12(1.10,1.15) |
| Diabetes | No | 1.25(1.23,1.27) | 1.18(1.16,1.20) | 1.00(0.98,1.02) |
| Glaucoma | No | **0.97(0.97,0.98)** | 1.01(0.99,1.02) | **0.97(0.95,0.99)** |
| Hip/Pelvic Fracture | No | 1.29(1.28,1.31) | 1.09(1.07,1.11) | 1.14(1.11,1.17) |
| Ischemic Heart Disease | No | **0.84(0.83,0.85)** | **0.92(0.91,0.93)** | **0.90(0.88,0.92)** |
| Depression | No | 1.04(1.03,1.05) | **0.97(0.96,0.98)** | 0.99(0.97,1.01) |
| Alzheimer'S Disease Or Dementia | No | 2.04(2.01,2.07) | 1.35(1.32,1.37) | 2.29(2.22,2.35) |
| Osteoporosis | No | **0.83(0.82,0.83)** | **0.80(0.79,0.81)** | **0.86(0.84,0.88)** |
| Rheumatoid Arthritis/Osteoarthritis | No | **0.90(0.89,0.91)** | **0.82(0.81,0.83)** | **0.80(0.78,0.82)** |
| Stroke/Transient Ischemic Attack | No | **0.87(0.87,0.89)** | 1.02(1.00,1.03) | 1.01(0.99,1.04) |
| Breast Cancer | No | 1.01(0.99,1.02) | 1.00(0.98,1.03) | 0.98(0.95,1.02) |
| Colorectal Cancer | No | 1.02(1.00,1.03) | **0.95(0.93,0.97)** | 1.03(1.00,1.07) |
| Prostate Cancer | No | 1.01(1.00,1.03) | **0.97(0.95,1.00)** | 1.00(0.97,1.03) |
| Lung Cancer | No | **0.97(0.95,0.99)** | 1.00(0.97,1.04) | *1.57(1.50,1.64)* |
| Endometrial Cancer | No | *1.04(1.01,1.07)* | 1.02(0.98,1.07) | 1.01(0.94,1.08) |
| Anemia | No | 0.99(0.98,1.00) | **0.94(0.93,0.96)** | *1.06(1.03,1.09)* |
| Asthma | No | **0.82(0.81,0.83)** | **0.88(0.87,0.90)** | **0.83(0.81,0.85)** |
| Hyperlipidemia | No | *1.07(1.06,1.09)* | **0.94(0.92,0.96)** | **0.77(0.75,0.79)** |
| Hyperplasia | No | **0.97(0.96,0.99)** | **0.88(0.86,0.90)** | **0.93(0.90,0.96)** |
| Hypertension | No | *1.78(1.72,1.85)* | *2.00(1.90,2.11)* | **0.80(0.74,0.85)** |
| Hypothyroidism | No | *1.01(1.00,1.02)* | **0.88(0.87,0.89)** | **0.93(0.91,0.95)** |
| ADHD And Other Conduct Disorders | No | *1.18(1.16,1.20)* | 0.99(0.97,1.01) | *1.14(1.11,1.18)* |
| Alcohol Use Disorders | No | *1.07(1.06,1.09)* | *1.04(1.01,1.06)* | 1.00(0.97,1.03) |
| Anxiety Disorders | No | *1.04(1.03,1.05)* | 0.99(0.98,1.00) | *1.08(1.06,1.10)* |
| Bipolar Disorder | No | *1.14(1.13,1.16)* | *1.10(1.08,1.12)* | *1.07(1.05,1.10)* |
| Traumatic Brain Injury | No | *1.08(1.06,1.10)* | *1.05(1.03,1.08)* | 1.02(0.99,1.07) |
| Drug Use Disorder | No | **0.90(0.89,0.92)** | *1.03(1.01,1.05)* | **0.94(0.91,0.97)** |
| Intellectual Disabilities | No | **0.85(0.83,0.87)** | **0.92(0.89,0.95)** | 1.00(0.94,1.06) |
| Learning Disabilities | No | 1.18(1.15,1.22) | 1.01(0.97,1.05) | *1.09(1.02,1.15)* |
| Other Developmental Delays | No | **0.65(0.62,0.68)** | **0.85(0.80,0.91)** | 0.93(0.84,1.04) |
| Personality Disorders | No | *1.05(1.03,1.06)* | *1.06(1.04,1.09)* | 0.99(0.96,1.02) |
| Schizophrenia | No | *1.51(1.49,1.53)* | *1.04(1.02,1.06)* | *1.20(1.17,1.23)* |
| Post-Traumatic Stress Disorder | No | **0.79(0.77,0.81)** | **0.92(0.89,0.96)** | **0.87(0.81,0.92)** |
| Cerebral Palsy | No | 1.02(0.98,1.06) | **0.93(0.88,0.98)** | 0.99(0.90,1.08) |
| Epilepsy | No | *1.02(1.01,1.04)* | **0.93(0.92,0.95)** | 1.01(0.99,1.04) |
| Cystic Fibrosis | No | *1.09(1.07,1.10)* | **0.97(0.95,0.99)** | 1.02(0.99,1.05) |
| Fibromyalgia, Chronic Pain And Fatigue | No | **0.87(0.87,0.88)** | **0.88(0.87,0.89)** | **0.82(0.81,0.84)** |
| Viral Hepatitis (General) | No | *1.04(1.02,1.06)* | 1.00(0.97,1.03) | 1.02(0.98,1.07) |
| Liver Disease Cirrhosis | No | **0.92(0.91,0.93)** | **0.95(0.93,0.96)** | *1.04(1.02,1.06)* |
| Hiv/Aids | No | *1.13(1.08,1.19)* | 0.94(0.88,1.00) | 0.96(0.86,1.06) |
| Leukemias And Lymphomas | No | **0.93(0.91,0.95)** | 1.02(0.99,1.05) | *1.22(1.18,1.27)* |
| Migraine And Other Chronic Headache | No | **0.77(0.76,0.78)** | **0.82(0.80,0.83)** | **0.77(0.75,0.79)** |
| Mobility Impairments | No | *1.04(1.02,1.05)* | 0.98(0.97,1.00) | 1.01(0.98,1.03) |
| Multiple Sclerosis And Transverse Myelitis | No | 1.02(0.99,1.05) | *1.16(1.11,1.21)* | 1.04(0.97,1.11) |
| Obesity | No | 1.02(1.01,1.03) | *1.16(1.14,1.17)* | **0.88(0.87,0.90)** |
| Overarching Opioid Use Disorder | No | **0.90(0.88,0.91)** | **0.94(0.92,0.96)** | **0.94(0.90,0.98)** |
| Peripheral Vascular Disease | No | *1.15(1.14,1.16)* | *1.05(1.04,1.07)* | *1.05(1.03,1.08)* |
| Spinal Cord Injury | No | *1.10(1.08,1.12)* | *1.07(1.05,1.09)* | *1.10(1.06,1.13)* |
| Tobacco Use Disorders | No | **0.98(0.97,0.99)** | *1.07(1.06,1.09)* | *1.08(1.05,1.10)* |
| Pressure Ulcers And Chronic Ulcers | No | *1.10(1.09,1.12)* | **0.98(0.96,0.99)** | *1.40(1.37,1.43)* |
| Deafness And Hearing Impairment | No | **0.95(0.95,0.96)** | **0.96(0.95,0.97)** | **0.98(0.96,1.00)** |
| Blindness And Visual Impairment | No | *1.16(1.14,1.18)* | 1.02(1.00,1.05) | *1.12(1.08,1.16)* |

**Bold type:** significant protective effect; *italics:* significant harmful effect

### 3.4 Risk of COVID-19 hospitalization

Overall, 142,004 (39.6%) COVID-19 patients were hospitalized for COVID-19. Current users of ACEI, ARB, direct factor Xa inhibitor and famotidine were associated with 2-9% decreased risk of COVID-19 hospitalization when compared with never users (Table 3). Hydroxychloroquine did not show any effect when recent starters were censored. The risk of a COVID-19 hospitalization increased monotonically with age (Table 4). Compared to the 65-69 age group, the risk increased by 11%, 43%, 66% and 97% for groups 70-74, 75-79, 80-84 and ≥85 respectively. Females enjoyed a 21% reduced risk compared to males. The risks of COVID-19 hospitalization for Asians and Hispanics were 5% and 15% higher than whites respectively. Poorer patients were associated with higher risks of hospitalization with COVID-19. The three co-morbidities associated with largest increase in risk were hypertension (100%), dementia (35%) and chronic kidney disease (33%).

### 3.5 Risk of mortality

Overall, 61,778 (17.2%) COVID-19 patients died. Current users of ACEI, ARB, warfarin, direct Xa inhibitor and clopidogrel among COVID-19 patients were all associated with lower risks of mortality, from 8% less than never users (clopidogrel) to 19% less (direct factor Xa inhibitor) (Table 3). Famotidine and hydroxychloroquine (with censoring of recent starters) showed no significant effect. Mortality risk increased monotonically with age (Table 4). Compared to patients 65-69, all other age groups exhibited increased risk of death: 12%, 55%, 98% and 212% increased risk among 70-74, 75-79, 80-84 and ≥85 respectively. All race groups were associated with notably greater mortality risk than whites. Unlike analyses of other outcomes, the poorest patients (dual-eligible) were associated with 28% decreased risk of death, while the second poorest patients (non-dual LIS) had 215% increased risk of death compared to patients not dually eligible and not receiving LIS. The three co-morbidities associated with the greatest mortality risk were dementia (129%) chronic kidney disease (58%) and lung cancer (57%).

## 4. Discussion

The most important finding of our study is that after controlling for age, gender, race, socio-economic, geographic factors and co-morbidities, there was an associated decline in mortality risk of 15% or greater among current users for ACEIs, ARBs and anticoagulants, and by a lesser amount (8%) among clopidogrel users. To the best of our knowledge, ours is first single source study to show such beneficial effects, though it has been hinted at in meta-analyses. [10, 11] One possible explanation for the benefit is that SARS-CoV-2 down-regulates ACE2 expression, resulting in unabated angiotensin II activity that may be responsible for organ damage in COVID-19. [6] ACEI and ARB reduce angiotensin II activity and can therefore be protective. Our findings provide strong support to the advice from professional societies and the WHO that ACEI and ARB be continued in COVID-19 patients. [40, 41] The benefits of antithrombotics can be explained by reduction in the thromboembolism observed in COVID-19 patients. [17, 19] These results strongly support the current advice about the use of anti-clotting drugs in COVID-19 patients. In addition, our study gives justification for the need of clinical trials to study the benefits of initiating ACEI or ARB treatment for patients with COVID-19.

The main strength of this study is the size of the study population and relative completeness of demographic, prescription and co-morbidity data. The U.S. has the largest collection of COVID-19 cases in the world. The elderly represents the most at-risk population for severe COVID-19. Our study population covered over 370,000 COVID-19 cases and 65,000 mortalities in U.S. patients over 65. According to CDC statistics, over 80% of COVID-19 mortalities occurred in patients over 65. A large U.K. study based on NHS records registered 10,926 COVID-19 deaths, our study surpassed that number by almost six-fold. [42] In terms of study drug usage among COVID-19 patients, our numbers were also much higher. For instance, one of the biggest original ACEI studies was from Italy, covering 1,502 ACEI users among COVID-19 patients. [7] We had 65 times that number. Sample sizes of this magnitude are not seen in other studies, including meta-analyses. [8, 10, 11] Moreover, other studies tended to be restricted to institutional or regional populations. Our Medicare population includes most US elderly individuals. By one estimate 93% of all US adults over 65 are enrolled to Medicare. [43] Unlike meta-analysis that pools multiple, not necessarily comparable data sources, we used a single data source that has relatively complete and longitudinal medication information with well-documented data capture procedures.

The emergence of hydroxychloroquine as a potential “game-changer” in COVID-19 had been accompanied by the dramatic rise in its use, not only among COVID-19 patients but in the general population as well. Even after the emergency use authorization was revoked by FDA, the use of hydroxychloroquine remained somewhat higher than the baseline level before the pandemic. This shows that media claims, even those that are eventually shown to be unsubstantiated or disproved, may have lasting impact on the public psyche. In our observational study, the use of hydroxychloroquine is a classic example of indication bias – the indication for the exposure is directly related to the outcome being observed. [44] Since we have longitudinal prescription data (starting from 2019), we were able to compensate for this by censoring patients who were first started on hydroxychloroquine during the pandemic. Indeed, our results show that, without censoring, the use of hydroxychloroquine would have appeared to be associated with an increase in the risk of COVID-19 diagnosis and death, but the effects disappeared with proper censoring.

Regarding the risk of catching COVID-19, we found that ACEI, ARB and antithrombotics were associated with a reduction in the risk of getting a COVID-19 diagnosis. One possible explanation is that if these drugs blunted the effects of COVID-19 infection, infected patients might have milder symptoms and would be more likely to not seek medical care and remain undiagnosed. The apparent reduction in risk of catching COVID-19 in patients over 75 could be related to the reduced level of social activity at extreme age, thus lowering the risk of exposure.

Apart from drug usage, we found the following risk factors for severe COVID-19 - advanced age, male, non-white, and co-morbidities including chronic kidney diseases, dementia, hypertension, heart diseases, chronic obstructive pulmonary disease, chronic liver diseases and some malignancies. These findings concur with similar studies and are well-documented. In our study, the group with the lowest income (dual-eligible) was associated with a reduced risk of death. One possible explanation is that this group has better access to healthcare, since patients are eligible to both Medicaid and Medicare.

We recognize the following limitations in our study. Drug data from Medicare claims are incomplete for hospitals and nursing homes. We did not include COVID-19 patients before April because the COVID-19 specific diagnosis code was not in use. We used all-cause mortality because of the lack of an accurate cause of death in CMS data.

## 5. Conclusion

Analysis of more than 370,000 Medicare enrollees over 65 diagnosed with COVID-19 showed that the use of ACEI, ARB, warfarin, direct factor Xa inhibitor and clopidogrel was associated with a reduction in the risk of catching COVID-19. The same drugs were also associated with a reduction in the risk of dying for patients who contracted COVID-19. Hydroxychloroquine and famotidine were not associated with significant effects in these outcomes.

## Data Availability

The Medicare claims data are available through the CMS Virtual Research Data Center.

## Acknowledgements

We would like to thank the diligent and helpful staff at the VRDC and Research Data Assistance Center (ResDAC), without them this study would not be possible.

## Contributorship Statement

KWF, SHB and CJM conceived and designed the study. SHB, FB and ZZ performed the statistical analysis. KWF drafted the manuscript and all authors contributed substantially to its revision.

## Data Availability Statement

The Medicare claims data are available through the CMS Virtual Research Data Center.

---

**Supplementary Table 1.** Drug classes, clinical drugs and usage frequencies among all our study population (ACEI angiotensin-receptor blocker; ARB angiotensin-receptor blocker)

| Drug class | Clinical drug | Number of claims | % within class |
| --- | --- | --- | --- |
| ACEI | AMLODIPINE BESYLATE/BENAZEPRIL | 907,955 | 3.03% |
|  | BENAZEPRIL HCL | 1,509,400 | 5.05% |
|  | BENAZEPRIL/HYDROCHLOROTHIAZIDE | 31,077 | 0.10% |
|  | CAPTOPRIL | 41 | 0.00% |
|  | ENALAPRIL MALEATE | 1,610,911 | 5.38% |
|  | ENALAPRIL/HYDROCHLOROTHIAZIDE | 77,437 | 0.26% |
|  | FOSINOPRIL SODIUM | 165,927 | 0.55% |
|  | FOSINOPRIL/HYDROCHLOROTHIAZIDE | 9,167 | 0.03% |
|  | LISINOPRIL | 20,637,915 | 68.98% |
|  | LISINOPRIL/HYDROCHLOROTHIAZIDE | 3,246,361 | 10.85% |
|  | MOEXIPRIL HCL | 8,609 | 0.03% |
|  | MOEXIPRIL/HYDROCHLOROTHIAZIDE | 2 | 0.00% |
|  | PERINDOPRIL ARG/AMLODIPINE BES | 9 | 0.00% |
|  | PERINDOPRIL ERBUMINE | 10,028 | 0.03% |
|  | QUINAPRIL HCL | 519,135 | 1.74% |
|  | QUINAPRIL/HYDROCHLOROTHIAZIDE | 50,329 | 0.17% |
|  | RAMIPRIL | 1,079,221 | 3.61% |
|  | TRANDOLAPRIL | 49,446 | 0.17% |
|  | TRANDOLAPRIL/VERAPAMIL HCL | 3,996 | 0.01% |
| ARB | AMLODIPINE BES/OLMESARTAN MED | 117,425 | 0.48% |
|  | AMLODIPINE BESYLATE/VALSARTAN | 112,278 | 0.46% |
|  | AMLODIPINE/VALSARTAN/HCTHIAZID | 29,880 | 0.12% |
|  | AZILSARTAN MED/CHLORTHALIDONE | 42,817 | 0.18% |
|  | AZILSARTAN MEDOXOMIL | 54,277 | 0.22% |
|  | CANDESARTAN CILEXETIL | 246,061 | 1.01% |
|  | CANDESARTAN/HYDROCHLOROTHIAZID | 59,711 | 0.25% |
|  | IRBESARTAN | 1,177,484 | 4.84% |
|  | IRBESARTAN/HYDROCHLOROTHIAZIDE | 117,725 | 0.48% |
|  | LOSARTAN POTASSIUM | 15,740,296 | 64.72% |
|  | LOSARTAN/HYDROCHLOROTHIAZIDE | 2,786,140 | 11.46% |
|  | OLMESARTAN MEDOXOMIL | 1,133,630 | 4.66% |
|  | OLMESARTAN MED/AMLODIPINE/HCTZ | 2,988 | 0.01% |
|  | OLMESARTAN/AMLODIPIN/HCTHIAZID | 62,604 | 0.26% |
|  | OLMESARTAN/HYDROCHLOROTHIAZIDE | 451,088 | 1.85% |
|  | TELMISARTAN | 361,087 | 1.48% |
|  | TELMISARTAN/AMLODIPINE | 2,926 | 0.01% |
|  | TELMISARTAN/HYDROCHLOROTHIAZID | 135,302 | 0.56% |
|  | VALSARTAN | 1,020,925 | 4.20% |
|  | VALSARTAN/HYDROCHLOROTHIAZIDE | 667,148 | 2.74% |
| HYDROXYCHLOROQUINE | CHLOROQUINE PHOSPHATE | 2,678 | 0.29% |
|  | HYDROXYCHLOROQUINE SULFATE | 916,812 | 99.70% |
|  | PRIMAQUINE PHOSPHATE | 40 | 0.00% |
| FAMOTIDINE | FAMOTIDINE | 2,146,195 | 99.41% |
|  | FAMOTIDINE/CA CARB/MAG HYDROX | 70 | 0.00% |
|  | FAMOTIDINE/PF | 2,446 | 0.11% |
|  | IBUPROFEN/FAMOTIDINE | 10,178 | 0.47% |
| WARFARIN | WARFARIN SODIUM | 3,041,385 | 100.00% |
| CLOPIDOGREL | CLOPIDOGREL BISULFATE | 7,735,118 | 100.00% |
| DIRECT FACTOR Xa<br>INHIBITOR | BETRIXABAN MALEATE | 6 | 0.00% |
|  | EDOXABAN TOSYLATE | 8,955 | 0.23% |
|  | RIVAROXABAN | 3,806,962 | 99.77% |

**Supplementary figure 1.**
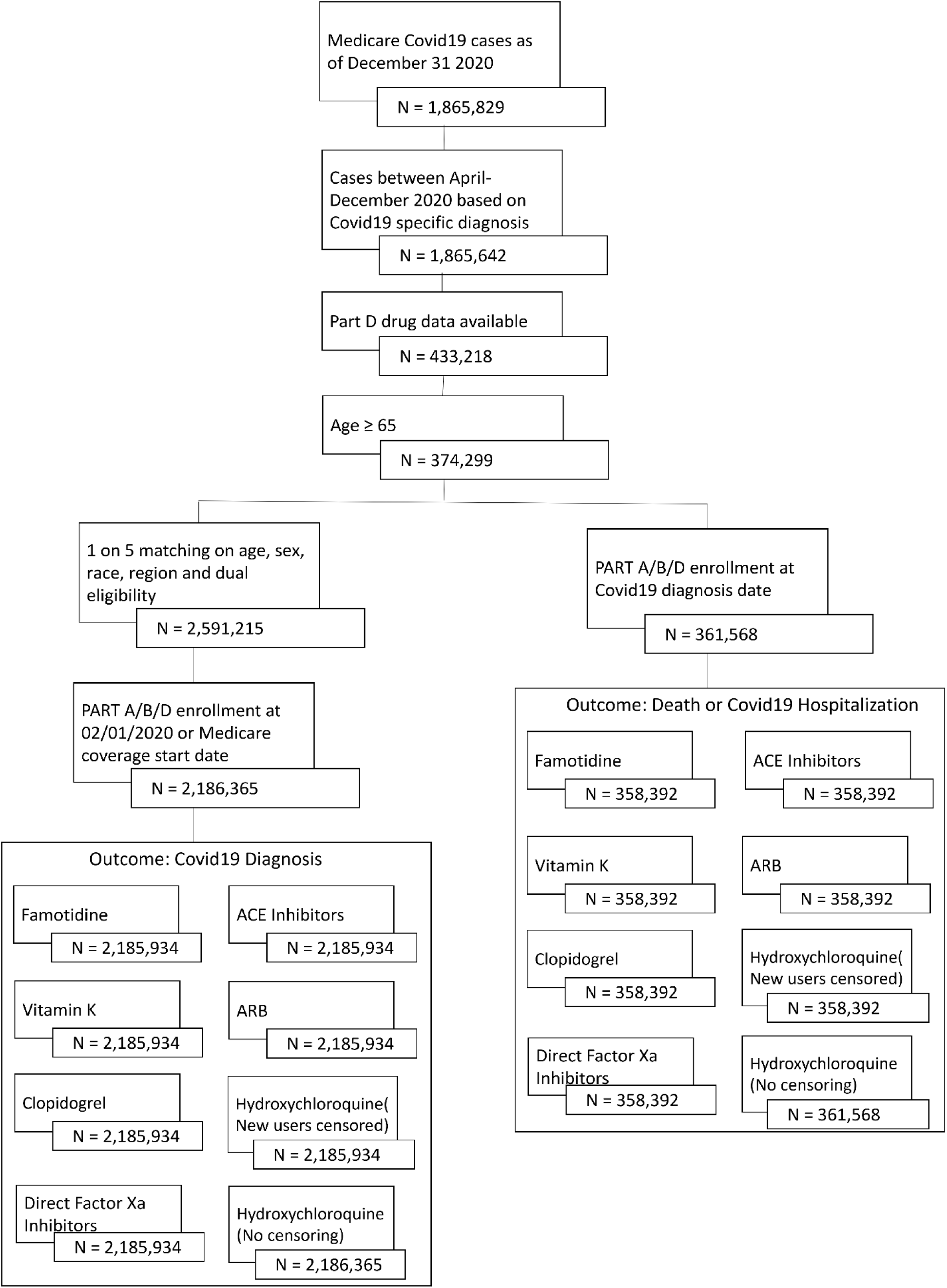
Consort diagram

**Supplementary Figure 2.**
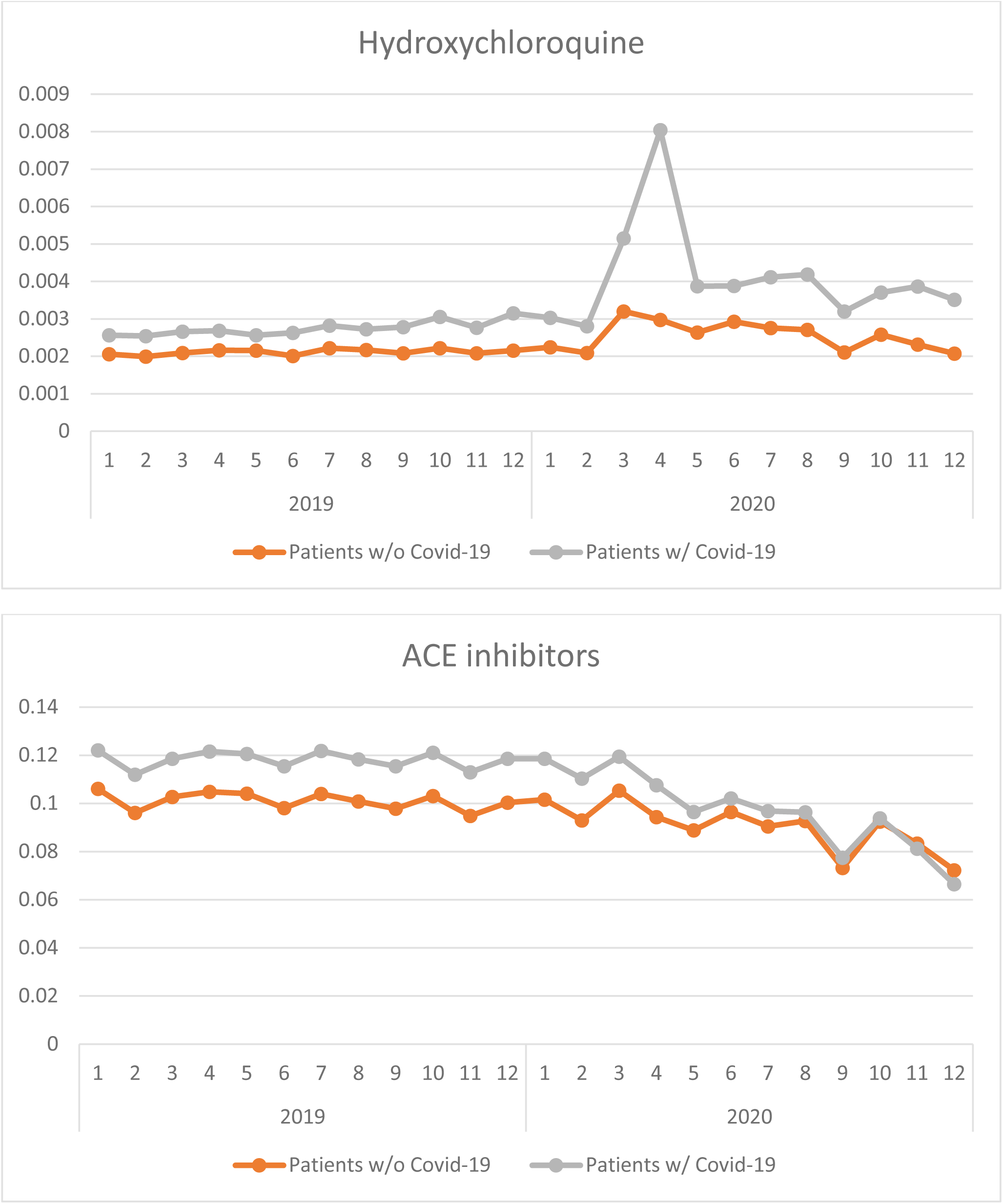
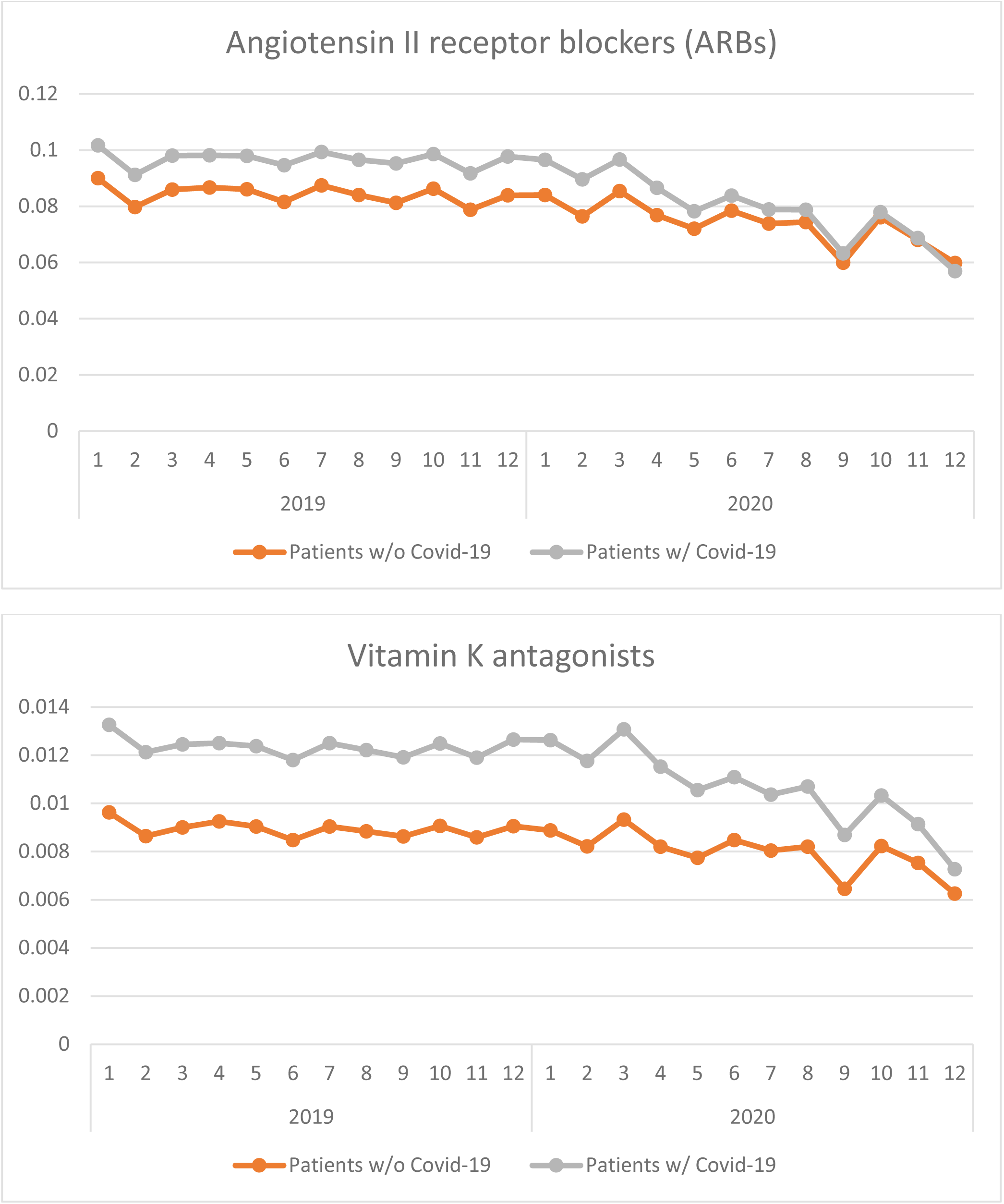
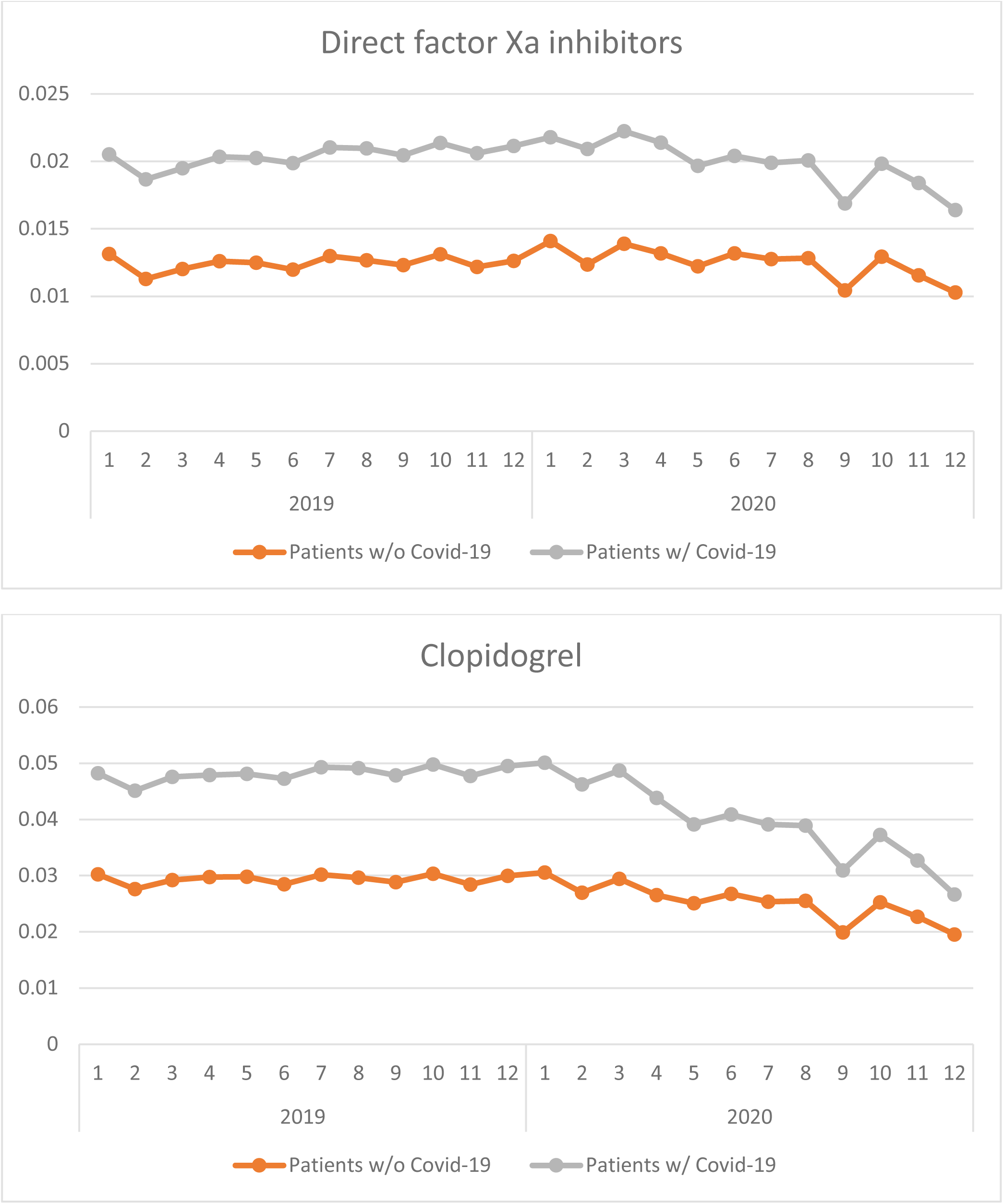
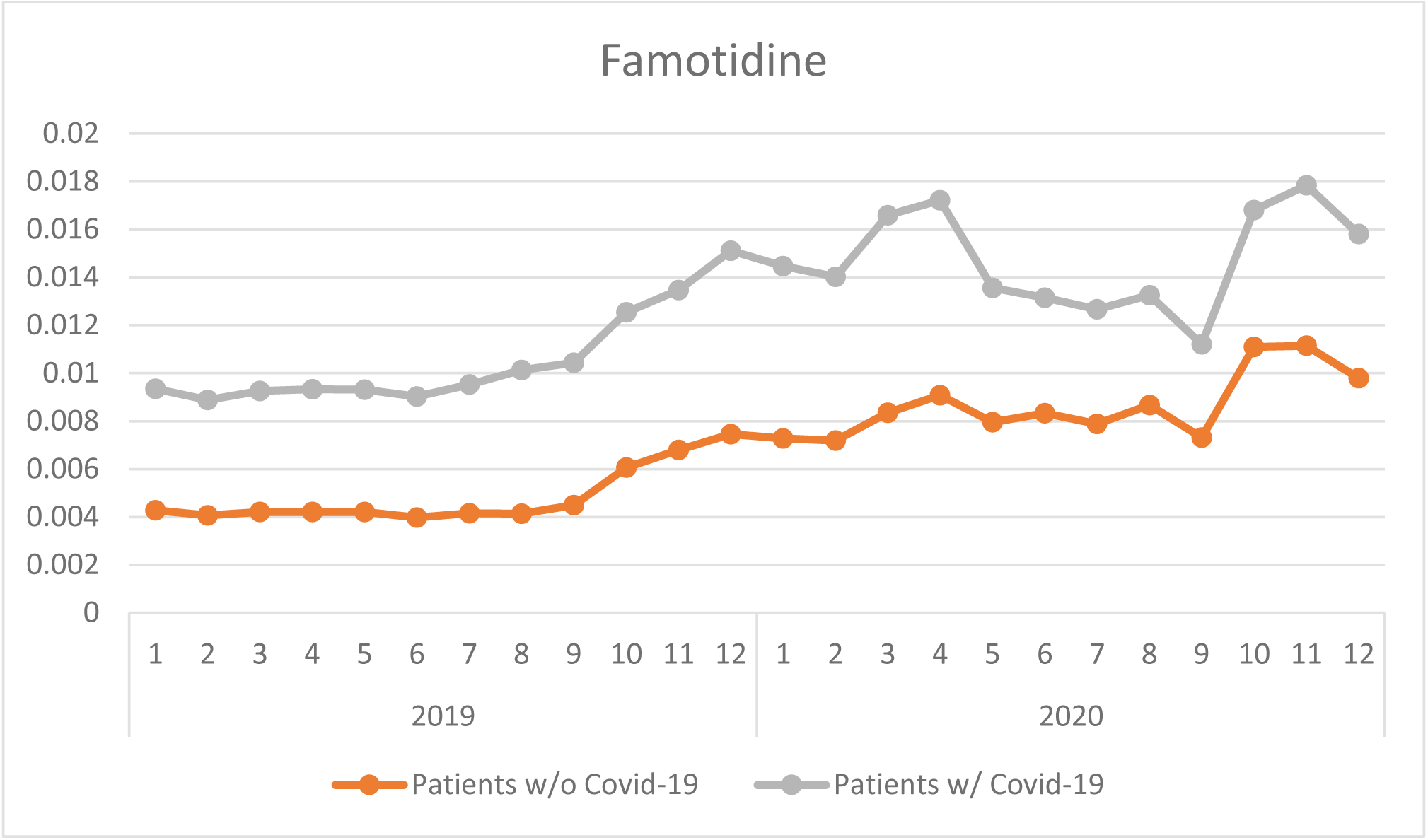
Trends of drug usage in 2019 and 2020 (w/o: without; w/: with)

## References

1. Ferrario, C.M., et al., Effect of angiotensin-converting enzyme inhibition and angiotensin II receptor blockers on cardiac angiotensin-converting enzyme 2. Circulation, 2005. 111(20): p. 2605–10.

2. Soler, M.J., et al., Pharmacologic modulation of ACE2 expression. Curr Hypertens Rep, 2008. 10(5): p. 410–4.

3. Li, W., et al., Angiotensin-converting enzyme 2 is a functional receptor for the SARS coronavirus. Nature, 2003. 426(6965): p. 450–4.

4. Hoffmann, M., et al., SARS-CoV-2 Cell Entry Depends on ACE2 and TMPRSS2 and Is Blocked by a Clinically Proven Protease Inhibitor. Cell, 2020. 181(2): p. 271–280 e8.

5. Li, J., et al., Association of Renin-Angiotensin System Inhibitors With Severity or Risk of Death in Patients With Hypertension Hospitalized for Coronavirus Disease 2019 (COVID-19) Infection in Wuhan, China. JAMA Cardiol, 2020. 5(7): p. 825–830.

6. Vaduganathan, M., et al., Renin-Angiotensin-Aldosterone System Inhibitors in Patients with Covid-19. N Engl J Med, 2020. 382(17): p. 1653–1659.

7. Mancia, G., et al., Renin-Angiotensin-Aldosterone System Blockers and the Risk of Covid-19. N Engl J Med, 2020. 382(25): p. 2431–2440.

8. Fosbol, E.L., et al., Association of Angiotensin-Converting Enzyme Inhibitor or Angiotensin Receptor Blocker Use With COVID-19 Diagnosis and Mortality. JAMA, 2020. 324(2): p. 168–177.

9. Reynolds, H.R., et al., Renin-Angiotensin-Aldosterone System Inhibitors and Risk of Covid-19. N Engl J Med, 2020. 382(25): p. 2441–2448.

10. Guo, X., Y. Zhu, and Y. Hong, Decreased Mortality of COVID-19 With Renin-Angiotensin-Aldosterone System Inhibitors Therapy in Patients With Hypertension: A Meta-Analysis. Hypertension, 2020. 76(2): p. e13–e14.

11. Barochiner, J. and R. Martinez, Use of inhibitors of the renin-angiotensin system in hypertensive patients and COVID-19 severity: A systematic review and meta-analysis. J Clin Pharm Ther, 2020.

12. Efficacy of Captopril in Covid-19 Patients With Severe Acute Respiratory Syndrome (SARS) CoV-2 Pneumonia (CAPTOCOVID) [clinical trial]. Available from: https://clinicaltrials.gov/ct2/show/NCT04355429.

13. Angiotensin Converting Enzyme Inhibitors in Treatment of Covid 19 [clinical trial]. Available from: https://clinicaltrials.gov/ct2/show/NCT04345406.

14. Valsartan for Prevention of Acute Respiratory Distress Syndrome in Hospitalized Patients With SARS-COV-2 (COVID-19) Infection Disease [clinical trial]. Available from: https://clinicaltrials.gov/ct2/show/NCT04335786.

15. Tang, N., et al., Abnormal coagulation parameters are associated with poor prognosis in patients with novel coronavirus pneumonia. J Thromb Haemost, 2020. 18(4): p. 844–847.

16. Bikdeli, B., et al., COVID-19 and Thrombotic or Thromboembolic Disease: Implications for Prevention, Antithrombotic Therapy, and Follow-Up: JACC State-of-the-Art Review. J Am Coll Cardiol, 2020. 75(23): p. 2950–2973.

17. Zhang, Y., et al., Coagulopathy and Antiphospholipid Antibodies in Patients with Covid-19. N Engl J Med, 2020. 382(17): p. e38.

18. Cui, S., et al., Prevalence of venous thromboembolism in patients with severe novel coronavirus pneumonia. J Thromb Haemost, 2020. 18(6): p. 1421–1424.

19. Klok, F.A., et al., Confirmation of the high cumulative incidence of thrombotic complications in critically ill ICU patients with COVID-19: An updated analysis. Thromb Res, 2020. 191: p. 148–150.

20. Rico-Mesa, J.S., et al., The Role of Anticoagulation in COVID-19-Induced Hypercoagulability. Curr Cardiol Rep, 2020. 22(7): p. 53.

21. Tremblay, D., et al., Impact of anticoagulation prior to COVID-19 infection: a propensity score-matched cohort study. Blood, 2020. 136(1): p. 144–147.

22. Russo, V., et al., Clinical impact of pre-admission antithrombotic therapy in hospitalized patients with COVID-19: A multicenter observational study. Pharmacol Res, 2020. 159: p. 104965.

23. Sivaloganathan, H., E.E. Ladikou, and T. Chevassut, COVID-19 mortality in patients on anticoagulants and antiplatelet agents. Br J Haematol, 2020.

24. Inama, G., et al., Coronavirus disease 2019 infection in patients with recent cardiac surgery: does chronic anticoagulant therapy have a protective effect? J Cardiovasc Med (Hagerstown), 2020. 21(10): p. 765–771.

25. Rossi, R., et al., Protective role of chronic treatment with direct oral anticoagulants in elderly patients affected by interstitial pneumonia in COVID-19 era. Eur J Intern Med, 2020. 77: p. 158–160.

26. Freedberg, D.E., et al., Famotidine Use Is Associated With Improved Clinical Outcomes in Hospitalized COVID-19 Patients: A Propensity Score Matched Retrospective Cohort Study. Gastroenterology, 2020. 159(3): p. 1129–1131 e3.

27. Bourinbaiar, A.S. and E.C. Fruhstorfer, The effect of histamine type 2 receptor antagonists on human immunodeficiency virus (HIV) replication: identification of a new class of antiviral agents. Life Sci, 1996. 59(23): p. PL 365–70.

28. Janowitz, T., et al., Famotidine use and quantitative symptom tracking for COVID-19 in non-hospitalised patients: a case series. Gut, 2020. 69(9): p. 1592–1597.

29. Keyaerts, E., et al., In vitro inhibition of severe acute respiratory syndrome coronavirus by chloroquine. Biochem Biophys Res Commun, 2004. 323(1): p. 264–8.

30. Vincent, M.J., et al., Chloroquine is a potent inhibitor of SARS coronavirus infection and spread. Virol J, 2005. 2: p. 69.

31. Sinha, N. and G. Balayla, Hydroxychloroquine and COVID-19. Postgrad Med J, 2020. 96(1139): p. 550–555.

32. Arshad, S., et al., Treatment with hydroxychloroquine, azithromycin, and combination in patients hospitalized with COVID-19. Int J Infect Dis, 2020. 97: p. 396–403.

33. Research Data Assistance Center (ResDAC). CMS Virtual Research Data Center (VRDC). Available from: https://www.resdac.org/cms-virtual-research-data-center-vrdc.

34. The Chronic Condition Warehouse. Chronic Conditions Data Warehouse: CCW Chronic Condition Algorithms.. Available from: https://www.ccwdata.org/web/guest/condition-categories.

35. Ernster, V.L., Nested case-control studies. Prev Med, 1994. 23(5): p. 587–90.

36. Azzato, E.M., et al., Prevalent cases in observational studies of cancer survival: do they bias hazard ratio estimates? Br J Cancer, 2009. 100(11): p. 1806–11.

37. Rosenbaum, P.R. and D.B. Rubin, The Central Role of the Propensity Score in Observational Studies for Causal Effects. Biometrika, 1983. 70: p. 41–55.

38. Wyss, R., et al., Use of Time-Dependent Propensity Scores to Adjust Hazard Ratio Estimates in Cohort Studies with Differential Depletion of Susceptibles. Epidemiology, 2020. 31(1): p. 82–89.

39. Centers for Disease Control and Prevention. COVID-19 Response. COVID-19 Case Surveillance Public Data Access, Summary, and Limitations. 2020; Available from: https://data.cdc.gov/Case-Surveillance/COVID-19-Case-Surveillance-Public-Use-Data/vbim-akqf.

40. Bozkurt, B., R. Kovacs, and B. Harrington, Joint HFSA/ACC/AHA Statement Addresses Concerns Re: Using RAAS Antagonists in COVID-19. J Card Fail, 2020. 26(5): p. 370.

41. World Health Organization. COVID-19 and the use of angiotensin-converting enzyme inhibitors and receptor blockers - Scientific Brief. 2020; Available from: https://www.who.int/news-room/commentaries/detail/covid-19-and-the-use-of-angiotensin-converting-enzyme-inhibitors-and-receptor-blockers.

42. Williamson, E.J., et al., Factors associated with COVID-19-related death using OpenSAFELY. Nature, 2020. 584(7821): p. 430–436.

43. Administration on Aging - U.S. Department of Health and Human Services. 2018 Profile of Older Americans. Available from: https://acl.gov/aging-and-disability-in-america/data-and-research/profile-older-americans.

44. Csizmadi, I., J.-P. Collet, and J.-F. Boivin, Bias and Confounding in Pharmacoepidemiology, in Pharmacoepidemiology. 2006. p. 791–809.

